# Mutational landscape of high-grade B-cell lymphoma with *MYC-, BCL2* and/or *BCL6* rearrangements characterized by whole-exome sequencing

**DOI:** 10.1101/2021.07.13.21260465

**Authors:** Axel Künstner, Hanno M. Witte, Jörg Riedl, Veronica Bernard, Stephanie Stölting, Hartmut Merz, Vito Olschewski, Wolfgang Peter, Julius Ketzer, Yannik Busch, Peter Trojok, Nikolas von Bubnoff, Hauke Busch, Alfred C. Feller, Niklas Gebauer

**Affiliations:** Medical Systems Biology Group, University of Lübeck, Ratzeburger Allee 160, 23538 Lübeck, Germany; Institute for Cardiogenetics, University of Lübeck, Ratzeburger Allee 160, 23538 Lübeck, Germany; University Cancer Center Schleswig-Holstein, University Hospital of Schleswig- Holstein, Campus Lübeck, 23538 Lübeck, Germany; Department of Hematology and Oncology, University Hospital of Schleswig-Holstein, Campus Lübeck, Ratzeburger Allee 160, 23538 Lübeck, Germany; Department of Hematology and Oncology, Federal Armed Forces Hospital Ulm, Oberer Eselsberg 40, 89081 Ulm; Hämatopathologie Lübeck, Reference Centre for Lymph Node Pathology and Hematopathology, Lübeck, Germany; HLA Typing Laboratory of the Stefan-Morsch-Foundation, 557565 Birkenfeld, Germany; Institut für Tranfusionsmedizin, Universitätsklinikum Köln. Kerpenerstr. 62, 50937 Köln; Department of Paediatrics, University Hospital of Schleswig-Holstein, Campus Luebeck, 23538 Luebeck, Germany

**Keywords:** High-grade B-cell lymphoma, mutational landscape, whole-exome sequencing

## Abstract

High-grade B-cell lymphoma accompanied with *MYC* and *BCL2* and/or *BCL6* rearrangements (HGBL-DH/TH) poses a cytogenetically-defined provisional entity among aggressive B-cell lymphomas that is traditionally associated with unfavorable prognosis.

To better understand the mutational and molecular landscape of HGBL-DH/TH we here performed whole-exome sequencing and deep panel next-generation-sequencing (NGS) of 47 clinically annotated cases. Oncogenic drivers, mutational signatures and perturbed pathways were compared with data from follicular lymphoma (FL), diffuse large-B-cell lymphoma (DLBCL) and Burkitt lymphoma (BL).

We find an accumulation of oncogenic mutations in NOTCH, IL6/JAK/STAT and NFκB signaling pathways and delineate the mutational relationship within the continuum between FL/DLBCL, HGBL-DH/TH and BL. Further, we provide evidence of a molecular divergence between *BCL2* and *BCL6* rearranged HGBL-DH. Beyond a significant congruency with the C3/EZB DLBCL cluster in *BCL2* rearranged cases on an exome-wide level, we observe an enrichment of the SBS6 mutation signature in *BCL6* rearranged cases. Differential gene set enrichment and subsequent network propagation analysis according to cytogenetically defined subgroups revealed an impairment of TP53 and MYC pathway signaling in *BCL2* rearranged cases, whereas *BCL6* rearranged cases lacked this enrichment, but instead exhibited showed impairment of E2F targets. Intriguingly, HGBL-TH displayed intermediate mutational features in all three aspects.

This study elucidates a recurrent pattern of mutational events driving FL into *MYC*-driven *BCL2* rearranged HGBL, unveiling the mutational pathogenesis of this provisional entity. Through this refinement of the molecular taxonomy for aggressive, germinal-center derived B-cell lymphomas, this calls into question the current WHO classification system, especially regarding the status of *MYC*/*BCL6* rearranged HGBL.

## Introduction

High-grade B-cell lymphoma with *MYC*-, *BCL2* and/or *BCL6* (HGBL) rearrangements poses a novel, yet provisional, cytogenetically-defined entity within the current WHO classification of lymphoid tumors. It is presumably allocated in the pathobiological continuum between diffuse large B-cell (DLBCL) and Burkitt lymphoma (BL) (1). The t(8;14)(q24;q32) IgH/*MYC* rearrangement constitutes the molecular hallmark of BL. This or further derivative chromosomal rearrangements that juxtapose *MYC* to a genomic enhancer, occur in approximately 10% of DLBCL and have been shown to correlate with inferior clinical outcome (2). The rearrangement is a driver of oncogenesis that is in approx. 50% of cases accompanied by additional rearrangements involving *BCL2* and/or *BCL6 and* referred to as double-hit (DH) or triple-hit (TH) lymphomas (3-11).

While the clinical outcome in HGBL-DH/TH patients is generally poor, recent studies have emphasized the significant impact of *MYC* translocation partners and defined *MYC*/*Ig*-rearrangements to be the most reliable predictors of adverse outcome (2, 7).

In a prior study, we discovered an elevated frequency of *TP53* impairment in *MYC*-driven DH/TH, whose presence was subsequently demonstrated for a subset of patients with a single-hit *MYC* translocation as well, indicating inferior outcome (12, 13).

By conventional cytogenetics HGBL-DH/TH were shown to recurrently harbor a complex karyotype (4). Data on the genetic basis of this entity, however, remains elusive. Several preliminary studies, predominantly focusing on HGBL-DH/TH with DLBCL morphology, have employed a panel-based next generation sequencing (NGS) approach (14-16). The insights from these studies were all restricted by gene-panel design and the associated clinicopathological data, yet their central assertions included a significant enrichment in mutations affecting *CREBBP, BCL2* and *KMT2D* alongside an overall reflection of the phenotypical gray-zone between DLBCL and BL. Most recently, Cucco *et al*. elucidated significant aspects of the molecular signature of HGBL in a panel-based sequencing and gene expression study, employing a 70-gene HaloPlex panel and an array-based gene expression approach. The authors restricted their study to samples with DLBCL morphology that stemmed from a clinical trial and the UK’s population-based Haematological Malignancy Research Network (17).

A comprehensive, exome-wide assessment of oncogenic driver mutations in HGBL-DH/TH, including cases with BL-like morphology is, however, still warranted and of vital importance to the refinement of the pathogenetic understanding of this clinically challenging entity ultimately enabling targeted therapeutic approaches.

We therefore conducted a whole-exome sequencing (WES) study on a large cohort of HGBL-DH/TH, validated by panel-based NGS and supplemented these data with a comprehensive clinicopathological assessment of the study group. Here we report on oncogenic drivers, somatic copy number alterations (SCNAs) and putative pathway perturbations, thus refining the molecular taxonomy of *MYC*-driven germinal-center-derived aggressive lymphomas.

## Materials and methods

### Case selection and clinicopathological characteristics

In a retrospective approach, we reviewed our institutional database to identify HGBL patients whose primary diagnostic biopsy specimen had been referred to the Reference center for Hematopathology University Hospital Schleswig Holstein Campus Lübeck and Hämatopathologie Lübeck for centralized histopathological panel evaluation between January 2007 and December 2019. For additional Information on clinicopathological work-up, please see **Supplementary materials and methods** and **Supplementary Table 1**.

This retrospective study was approved by the ethics committee of the University of Lübeck (reference-no 18-356) and conducted in accordance with the declaration of Helsinki. Patients had given written informed consent regarding routine diagnostic and academic assessment of their biopsy specimen including molecular studies at the Reference center for Hematopathology and transfer of their clinical data.

### Whole exome and targeted amplicon-based sequencing

Whole exome sequencing of n = 47 HGBL-DH/TH samples was performed by a hybrid capture approach with the Agilent SureSelect Human All Exon V6 library preparation kit (Agilent Technologies) followed by Illumina short read sequencing on a NovaSeq platform (Illumina) to an average depth of 303x (standard deviation ±152x; median 237x) by Novogene (UK) Co., Ltd. Seeking to validate the initial delineation of the exome sequencing-derived mutational landscape in HGBL we employed our in-house custom AmpliSeq panel (Thermo Fisher Scientific, Waltham, Massachusetts, USA) for targeted amplicon sequencing (tNGS), encompassing all coding exons of 43 genes (see **Supplementary Table 2**) in 21 cases. Raw paired-end data (*fastq* format) was trimmed and quality filtered using FASTP (v0.20.0; minimum length 50bp, max. unqualified bases 30%, trim tail set to 1) (18) and trimmed reads were mapped to GRCh37/hg19 using BWA MEM (v0.7.15) (19). Resulting alignment files in *SAM* format were cleaned, sorted, and converted into *BAM* format using PICARD TOOLS (v2.18.4). Single nucleotide variants (SNVs), as well as short insertions and deletions (InDels) were identified following the best practices for somatic mutations calling provided by GATK (20). Somatic copy number aberrations (SCNAs) were identified by CONTROL-FREEC (v11.4) (21). For further details on nucleic acid extraction, panel sequencing, single nucleotide and copy number variant calling please see **Supplementary methods**.

### Mutational deleteriousness and significance, network propagation, gene set enrichment and mutational cluster analysis

The MUTSIGCV algorithm was employed on WES data to delineate significantly mutated genes within the study cohort while deleteriousness was assessed via the CADD v1.3. The acquired genomic data were then processed through the LymphGen algorithm and underwent manual screening for an enrichment in overlapping aberrations with the molecular clusters proposed by Chapuy *et al*. followed by validation through a logistic regression framework (22, 23). Cytogenetically defined subgroups (HGBL with *MYC* and *BCL2* aberrations, HGBL with *MYC* and *BCL6* aberrations, and HGBL-TH) underwent differential downstream analysis by a network propagation approach simulating a protein-protein interaction network. Subsequently a gene set variation analysis was performed against HALLMARK gene sets. For details including statistical approaches correlating molecular and clinicopathological findings please see **Supplementary methods**.

### Data availability

Sequencing data in bam format from WES and panel sequencing have been deposited in the European genome-phenome archive (EGA) under the accession number EGAS00001005420.

## Results

### Clinicopathological characteristics of the study group

We collected 47 cases of HGBL-DH/TH at diagnosis with sufficient FFPE tissue samples for molecular studies (median age 71; range 35 – 89 years) all of which were included in the final analysis, following successful library preparation for WES. There was insufficient clinical follow-up in 9/47 (19%) cases. An underlying HIV infection was clinically excluded in all cases. The majority of patients in our study were male (25/47; 53%) and presented with advanced stage disease (24/38 stage III/IV; 63%) and an adverse prognostic constellation (24/38 (63%) R-IPI > 2). Most patients received an intensive CHOP-like therapeutic frontline approach (25/38; 66%). The overall response rate after first line (immuno-) chemotherapy was 76% resembling a general therapeutic response in 29/38 cases.

**Table 1** summarizes the baseline characteristics of all HGBL-DH/TH cases included in the current study. Histologically, the predominant morphology was that of DLBCL (NOS) (32/47), however, Burkitt-like morphology and immunophenotype was present in 15/47 patients. Cytogenetically 21/47 cases presented with *MYC*/*BCL2*, 17/47 with *MYC*/*BCL6* double-hit constellation and 9 triple-hit lymphomas were included. *MYC* translocation partner revealed *MYC*-Ig rearrangement in 8/17 cases. The treatment outcome in our cohort was unfavorable yet in keeping with previous data reported by Rosenwald and colleagues (2). For confirmatory purposes, we included four cases, which were assessed for *TP53* mutation status in a previous study and were able to validate both cytogenetic as well as molecular observations (12).

**Table 1.** Clinical characteristics of the study group.

| Characteristics | HGBL<br>(n = 47) | DHL-*BCL2*<br>(n = 21) | DHL-*BCL6*<br>(n = 17) | THL<br>(n = 9) |
| --- | --- | --- | --- | --- |
| Insufficient FU | 9 (19.1%) | 4 (19.0%) | 2 (11.8%) | 3 (33.3%) |
| Age<br>(yrs.; median + range) | 71 (35 – 89) | 73 (35 – 88) | 72 (35 – 89) | 66 (42 – 76) |
| Sex |  |  |  |  |
| Female | 22 (46.8%) | 8 (38.1%) | 11 (64.7%) | 3 (33.3%) |
| Male | 25 (53.2%) | 13 (61.9%) | 6 (35.3%) | 6 (66.7%) |
| **R-IPI** |  |  |  |  |
| 0 | 1 (2.6%) | 1 (5.9%) | - | - |
| 1-2 | 13 (34.2%) | 5 (29.4%) | 6 (40.0%) | 2 (33.3%) |
| >2 | 24 (63.2%) | 11 (64.7%) | 9 (60.0%) | 4 (66.7%) |
| **Stage (Ann Arbor)** |  |  |  |  |
| I | 5 (13.2%) | 2 (11.8%) | 3 (20.0%) | - |
| II | 9 (23.7%) | 4 (23.5%) | 3 (20.0%) | 2 (33.3%) |
| III | 5 (13.2%) | 1 (5.9%) | 2 (13.3%) | 2 (33.3%) |
| IV | 19 (50.0%) | 10 (58.8%) | 7 (46.7%) | 2 (33.3%) |
| **B-Symptoms** |  |  |  |  |
| Yes | 20 (52.6%) | 10 (58.8%) | 7 (46.7%) | 3 (50.0%) |
| No | 18 (47.4%) | 7 (41.2%) | 8 (53.3%) | 3 (50.0%) |
| **Extranodal sites** |  |  |  |  |
| 0 | 9 (23.7%) | 3 (17.6%) | 4 (26.7%) | 2 (33.3%) |
| 1-2 | 28 (73.7%) | 14 (82.4%) | 10 (66.7%) | 4 (66.7%) |
| >2 | 1 (2.6%) | - | 1 (6.7%) | - |
| **ECOG PS** |  |  |  |  |
| 0-1 | 20 (52.6%) | 8 (47.1%) | 8 (53.3%) | 4 (66.7%) |
| ≥2 | 18 (47.4%) | 9 (52.9%) | 7 (46.7%) | 2 (33.3%) |
| **LDH** |  |  |  |  |
| Normal | 7 (18.4%) | 3 (17.6%) | 3 (20.0%) | 1 (16.7%) |
| Elevated | 31 (81.6%) | 14 (82.4%) | 12 (80.0%) | 5 (83.3%) |
| **CNS involvement at diagnosis** |  |  |  |  |
| Yes | 2 (5.3%) | - | 2 (13.3%) | - |
| No | 36 (94.7%) | 17 (100.0%) | 13 (86.7%) | 6 (100.0%) |
| **Morphology** |  |  |  |  |
| DLBCL-like | 32 (68.1%) | 17 (80.9%) | 12 (70.6%) | 3 (33.3%) |
| Burkitt-like | 15 (31.9%) | 4 (19.0%) | 5 (29.4%) | 6 (66.7%) |
| **Frontline therapy regimen** |  |  |  |  |
| CHOP-like | 25 (65.8%) | 11 (64.7%) | 10 (66.7%) | 4 (66.7%) |
| R-based | 31 (81.6%) | 14 (82.4%) | 12 (80.0%) | 5 (83.3%) |
| Intensified* | 13 (34.2%) | 7 (41.1%) | 3 (20.0%) | 3 (50.0%) |
| Less intensive** | 9 (23.7%) | 4 (23.5%) | 4 (26.7%) | 1 (16.7%) |
| Refusal | 1 (2.6%) | - | 1 (6.7%) | - |
| CHOP, cyclophosphamide/hydroxadaunorubicin/vincristine/prednisolone; CNS, central nervous system; DHL, double-hit lymphoma; DLBCL, diffuse large B-Cell Lymphoma; ECOG, Eastern cooperative oncology group; FU, follow-up; HGBL, high grade B-cell lymphoma; LDH, Lactate dehydrogenase; PS, performance status; R, rituximab; R-IPI, revised International Prognostic Index; THL, triple-hit lymphoma; Yrs., years.<br>*Intensified regimens: B-ALL, GMALL, CHOEP (additional etoposide), EPOCH<br>**Less intensive regimens: Bendamustin, mini-CHOP (50% dose reduction), rituximab mono |  |  |  |  |

### The mutational landscape of HGBL-DH/TH identified by WES

To characterize the mutational landscape in an extensive cohort of HGBL-DH/TH cases, we successfully performed WES in 47 patient-derived tumor biopsies and matched constitutional DNA in seven cases. We further applied the analytical framework outlined above to analyze WES data in the absence of paired germline DNA in the majority of cases. Following the primary identification of SNVs and indels in individual samples and subsequent filtering to correct for FFPE-derived artefacts and spurious mutations, we applied the MutSig2CV algorithm and thereby identified 22 significant candidate driver genes (p < 0.001; 13 genes with q < 0.1; **Supplementary Table 3**) (24).

All HGBL-DH/TH cases carried mutations in genes of oncogenic relevance according to our bioinformatic annotations. In total, we described 10,092 presumably harmful somatic mutations (cut-off see materials and methods) involving 5,521 genes after variant filtering. Of these, SNVs and InDels represented 74.1% of the mutations (7,479 SNVs). Among them, missense mutations were the most frequent alterations (85.2%), followed by nonsense (5.7%) and InDels (5.6%), while splice-site mutations posed 3.3% of somatic mutations (**Figure 1A**). Displaying an overall intermediate tumor mutational burden (median 3.974; range 1.065 – 18.234 mutations/Mbase; **Figure 1B**), HGBL-DH/TH revealed no evidence of MSI-related hypermutations, which is in keeping with observations in DLBCL (0.3%), yet differs from other aggressive lymphomas (e.g., primary mediastinal B-cell lymphoma) (25).

**Figure 1.**
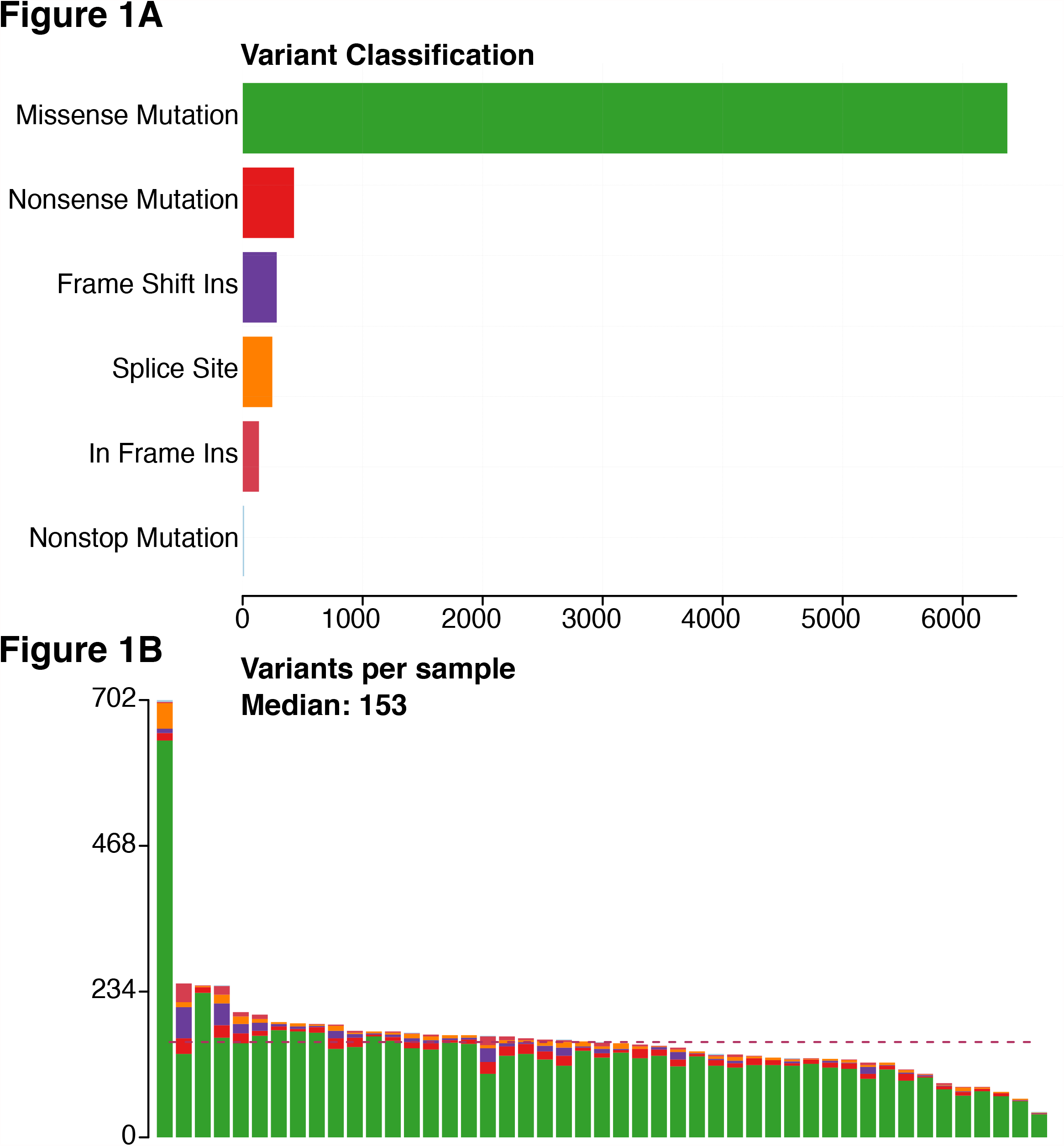
Panel (A) shows the number of variants stratified by variant classification while panel (B) delineates the number of mutations per sample with a median of 153 mutations per sample.

Upon comparative analysis of WES and targeted resequencing data, we were able to demonstrate a concordance rate of 92.0% (46/50 in 18 matched samples) of mutational calls, prompting high confidence in mutational calls derived from WES, even in non-germline-paired cases. A comprehensive description of all variants described by WES as well as panel based NGS is provided in **Supplementary Tables 4** and **5**. Nevertheless, we observed a significant enrichment of non-germline matched samples in non-synonymous SNVs, which prompted us to include significantly mutated genes according to the MUTSIGCV analysis, only. As an exception to this rule, we also included *MYC* mutations below the statistical significance level due to their previously established clinical and functional relevance.

### Recurrent copy number alterations in HGBL-DH/TH

We investigated our HGBL-DH/TH cohort for SCNVs employing the CONTROL-FREEC (21) algorithm in tumor-normal and tumor-only mode, respectively, followed by GISTIC2.0 (26) analysis. The analysis excluded chromosomes X and Y as well as common benign copy number variants defined by the Broad Institute’s panel of normal. Upon cross-referencing our findings with genomic loci of known oncogenes, tumor-suppressors and elements of significant signaling pathways we identified recurrent copy number gains in oncogenes such as *MEF2B* and *CSF1R*, which have previously been implicated in the pathogenesis of malignant lymphomas (27, 28). Further, copy number losses in tumor suppressors like *NPM1* were recurrently identified (**Figure 2A, B**). No significant differences were detected for genes affected by copy number alterations between the three cytogenetically defined subgroups (Fisher exact test p > 0.05 after Bonferroni correction for multiple testing). Common CNVs, as defined by the above referenced panel of normal were encountered at the expected frequencies (29, 30).

**Figure 2.**
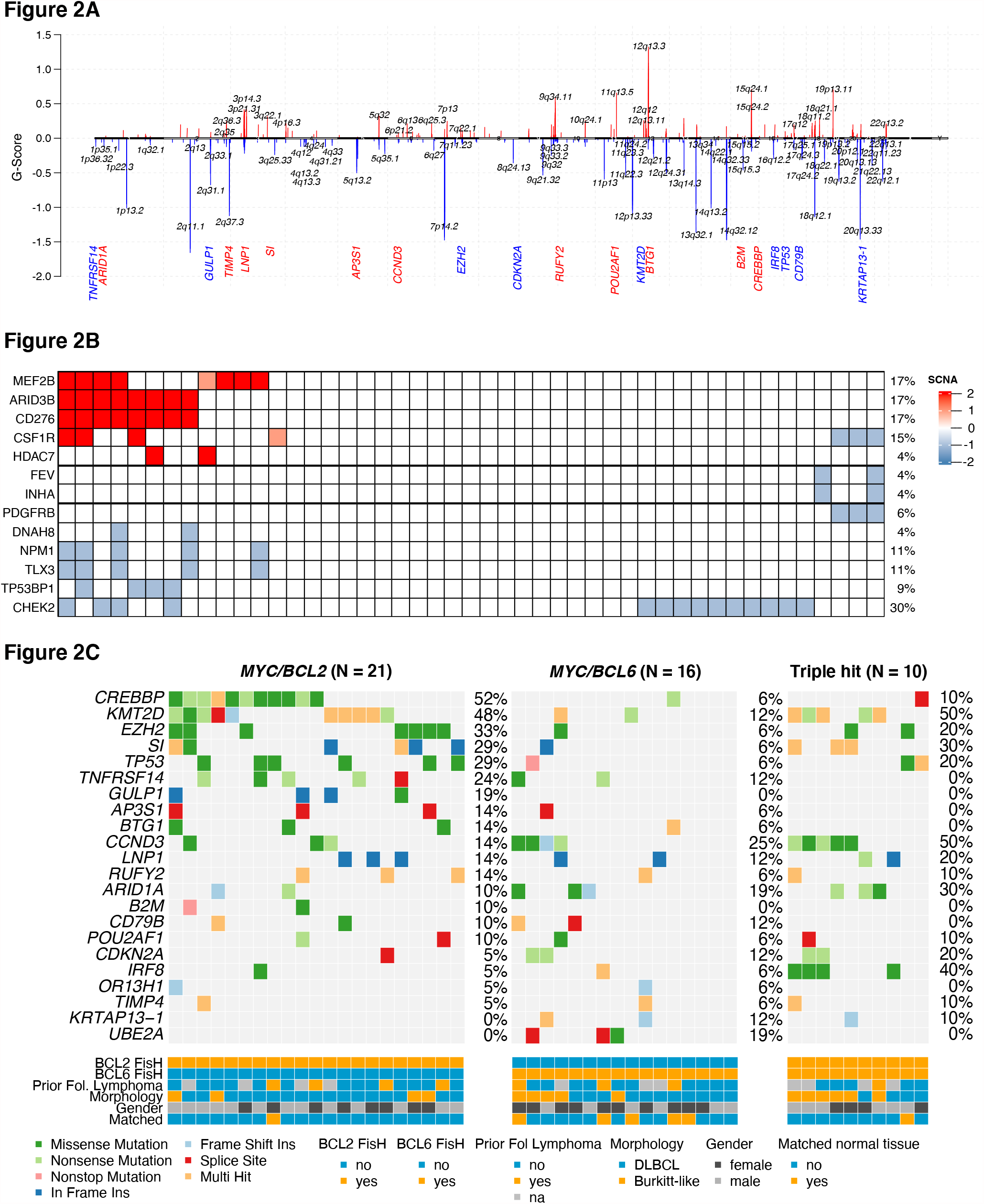
Co-oncoplot for genes identified as significant driver genes by MUTSIGCV (p < 0.001; n = 22) in our cohort stratified by cytogenetical subtypes is shown in Panel (A); different types of mutations are colour coded. Additionally, covariates are shown below the plot for each sample. (B) Location of SCNVs along the genome (red bars denote gains; blue bars denote losses; gene names refer to affected oncogenes and genes related to oncogenetic pathways). Panel (C) displays selected genes (oncogenes and genes implicated in oncogenetic pathways) affected by SCNVs.

### Significantly mutated candidate driver genes and mutational signatures

Putative candidate driver genes comprised several genes previously implicated in HGBL-DH/TH pathogenesis, such as *KMT2D, CREBBP* and *TP53* alongside several further mutated genes such as *CDKN2A, LNP1* or *SI* (14). Established mutational drivers known from other B-cell lymphoproliferative disorders (e.g., FL, DLBCL, BL) were recurrently encountered (e.g., *CCND3, ARID1A*) (**Figures 2C; 3A**) (31, 32). In our limited cohort, distinctions regarding subtype-specific mutational signatures were found to be marginal among *BCL2*/*BCL6* status or Burkitt-like vs non-Burkitt like morphology. However, we found *CCND3* mutations, previously reported as driver mutations in BL pathogenesis, to be significantly enriched in HGBL-DH/TH patients with Burkitt-like morphology (9/15 vs 3/28). This observation hints at partially similar molecular paths of pathogenesis between BL and HGBL with Burkitt-like morphology. Further, an enrichment of mutations affecting *CREBBP* in HGBL-DH/TH patients with *BCL2* rearrangement was observed, which is well in keeping with its proposed fundamental role in FL pathogenesis. Distribution of mutations within selected, significantly mutated genes is depicted in **Supplementary Figure 1**. Additional profiling of mutational signatures driving HGBL-DH/TH revealed a predominance of the SBS5 signature alongside the emphasized occurrence of the SBS6 signature (implicated in defective DNA mismatch repair) in patients with *BCL6* rearrangements (**Supplementary Figure 2, Supplementary Table 6**).

### Comparative analysis of mutational landscape in HGBL-DH/TH, related entities and molecular clusters in DLBCL

Next, we sought to refine the genomic taxonomy of aggressive germinal-center-derived B-cell lymphomas and to investigate the mutational commonalities and differences between HGBL-DT/TH and other related pathological entities. These were subsequently selected for their similar features of B-cell differentiation (ABC-type DLBCL, FL, GCB-type DLBCL and BL) and a comparative analysis of candidate mutational drivers in HGBL-DH/TH (as described earlier) and cBioPortal cohorts of ABC-type DLCBL (n = 67), GCB-type DLBCL (n = 45 (22)), BL (n = 108(33)) and FL (n = 199(33)) was conducted. Interestingly, we identified one overlapping candidate driver common to all entities (*KMT2D)*. Additionally, *CREBBP* was found in all entities except ABC-type DLCBL. Mutations affecting *EZH2, IRF8* and *TNFRSF14* were, however, specifically occured in HGBL-DT/TH and FL/GCB-type DLBCL, while *CCND3* mutations appeared to be a pathogenetic feature shared between BL and HGBL-DT/TH. Additionally, *TP53* mutations posed a predominant feature of aggressive lymphomas present in all HGBL-DH/TH, GCB-type DLBCL and BL types and therefore most likely acquired during high-grade transformation (**Figure 3G, H**). In basic accordance with previous studies, our data suggest a common origin especially for *BCL2* rearranged HGBL-DH/TH and FL/GCB-type DLBCL (14, 17).

**Figure 3.**
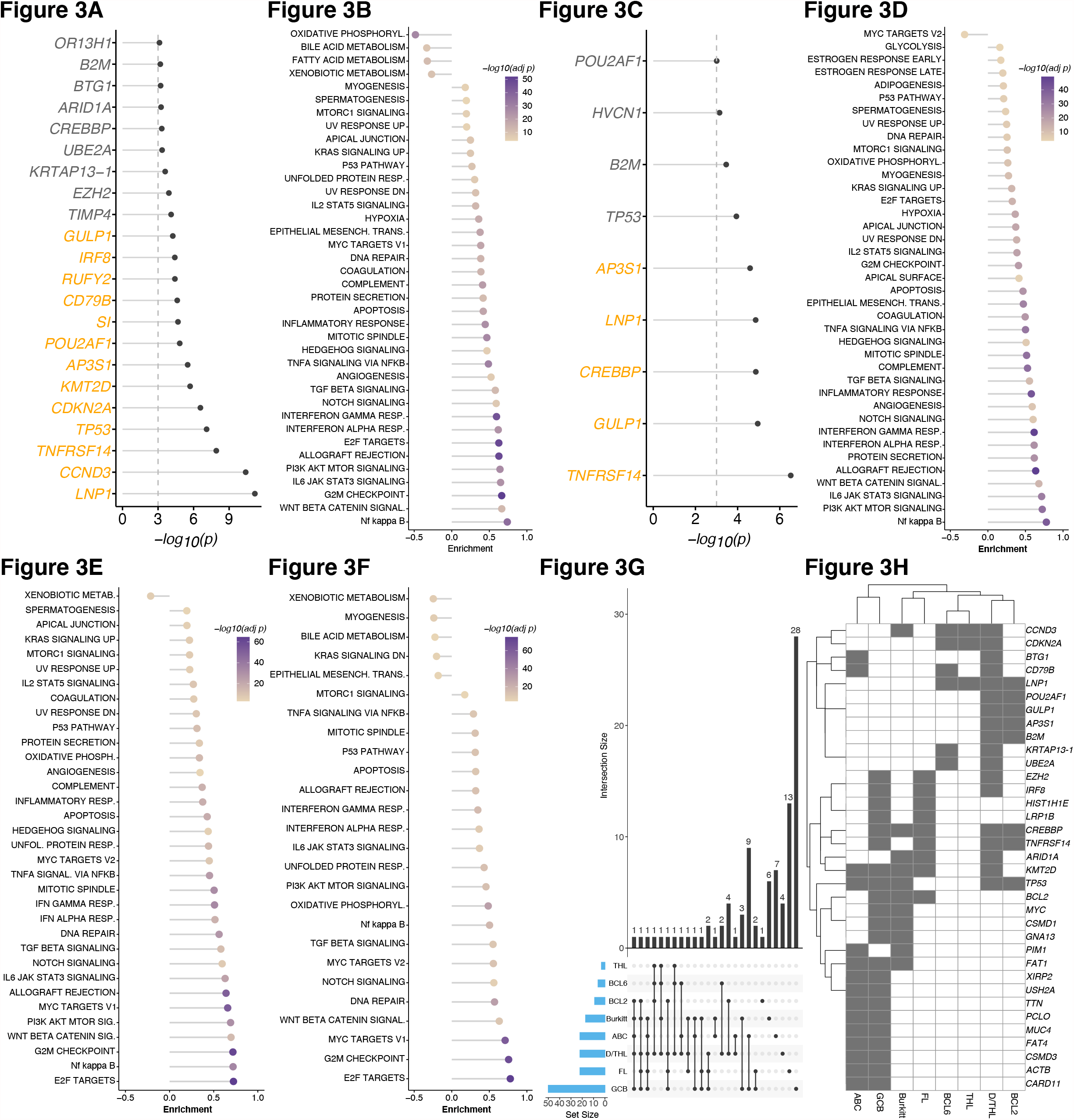
(A) Significance levels for all HGBL-DH/TH MUTSIGCV genes (p < 0.001; gene names in orange indicate q < 0.1) and (B) HALLMARK gene sets and NFκB pathway enrichment for network propagation analysis of significant MUTSIGCV genes (MUTSIGCV p < 0.001). (C) Significant levels for *MYC/BCL2* subgroup (MUTSIGCV p < 0.001) and (D) results for enrichment for network propagation analysis against HALLMARK gene sets including NFκB pathway. Panels (E) and (F) show enrichment results for cytogenetical subgroups *MYC/BCL6* and triple hit, respectively (*MYC/BCL6* genes included: CDKN2A, CD78B, LNP1, KRTAP13-1, UBE2A, CCND3; triple hit genes included: CDKN2A, LNP1, CCND3). UpSet plot (G) showing the overlap of MUTSIGCV genes using our HGBL-DH/TH (D/THL) cohort, the cytogenetical subgroups (BCL2, BCL6, THL), as well as cohorts of ABC-type DLBCL (n = 67(33)), GCB-type DLBCL (n = 45 (22)), BL (n = 108(38)) and FL (n = 199(38)) (all retrieved via cBioPortal); (H) shows the overlap between the five lymphoma subtypes and the three cytogenetical subtypes of HGBL-DH/TH.

Upon comparative investigation of our current data and mutational clusters, previously described in DLBCL, we observed a striking predominance of C3/EZB cluster cases in the *BCL2* rearranged subgroup according to the integrative molecular classification proposed by Chapuy *et al*. and Wright *et al*., respectively. This is in keeping with a significant enrichment of these cases with DLBCL morphology in terms of *MYC* rearrangement status (**Figure 4, Supplementary Figure 3**) (22, 23). Complementary to our analysis, employing the LymphGen algorithm (cf. **Supplementary Table 7**), a logistic regression indicated a significantly different number of mutated genes in C3 between the HGBL subtypes. HGBL harboring only *BCL2* were shown to exhibit the highest number of mutated C3 genes, while HGBL with *BCL6* alterations had the lowest number of mutated C3 genes (*BCL2/6* cases: (p = 6.059*10^−5^, adj R2 = 0.3108). In contrast to triple hit cases, HGBL with *MYC* and an isolated additional *BCL6* rearrangement showed a significant decrease in the number of mutated C3 genes (p = 2.00*10^−5^, estimate: -1.7589) (**Supplementary Figure 4, Supplementary Table 8**). Within the subgroup of *BCL6* rearranged cases, the BN2 cluster was more prominent than the EZB cluster. In keeping with their strong affinity towards the C3/EZB cluster, BCL2 rearranged cases exhibited an enrichment for mutations in *CREBBP* and *KMT2D*, while *BCL6* rearranged cases were, in contrast, enriched for mutations in *ARID1A*. The vast majority of triple-hit cases was also classified within the EZB cluster.

**Figure 4.**
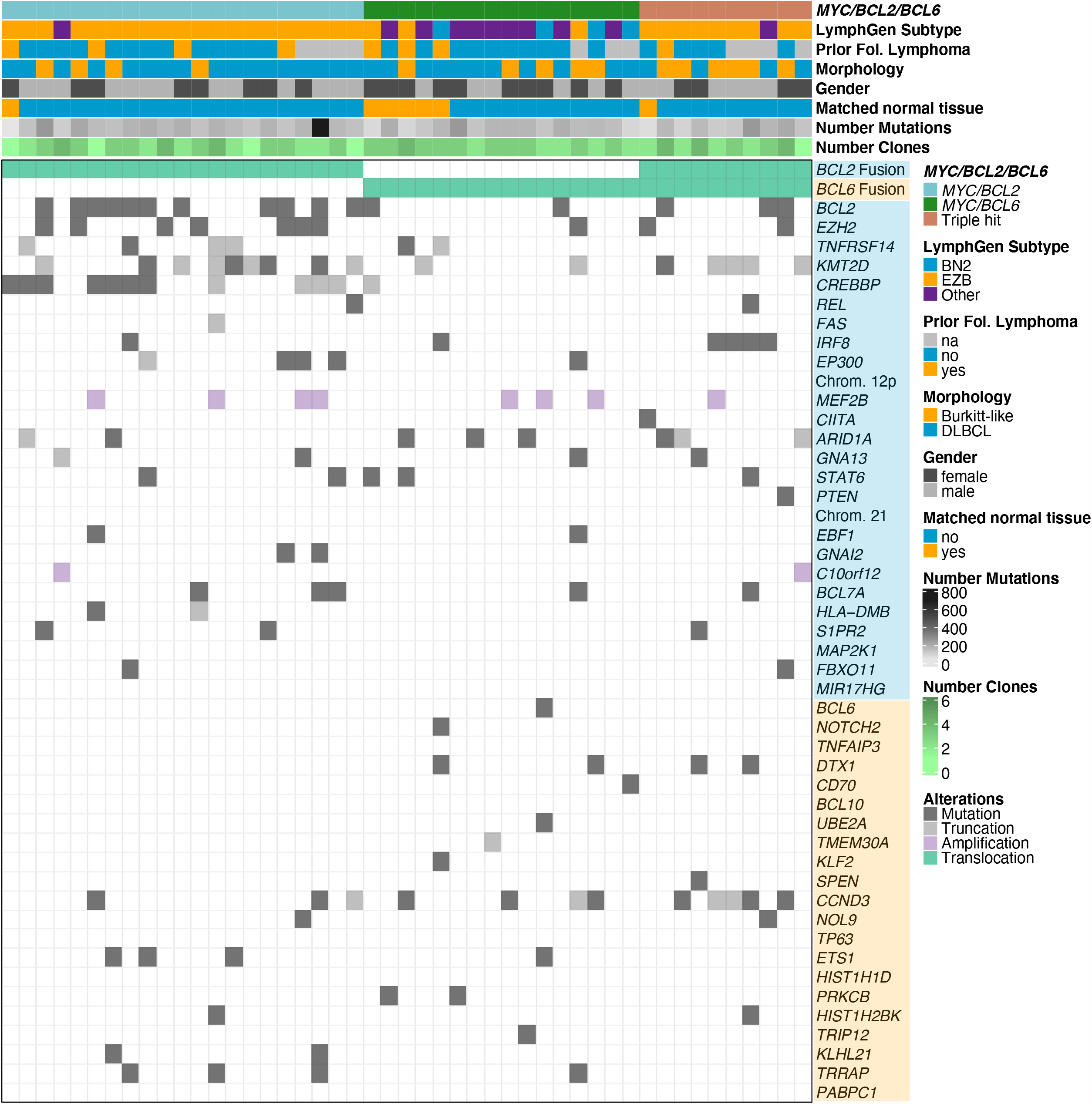
Allocation of HGBL-DH/TH samples unto the molecular subgroups/clusters of DLBCL, according to LymphGen based on their mutational signature. Additionally, covariates are shown above the plot for each sample. Row names refer to chromosomal alterations, genes and fusions and are background coloured by their specific subtype (blue = BN2, orange = EZB).

### Mutational impairment of NOTCH, RTK-RAS and TP53 signaling in HGBL-DH/TH

Cumulatively, we detected genetic lesions, putatively impairing NOTCH signaling in 74% of HGBL-DH/TH patients. Expanding on previously reported *CREBBP, EP300* and *DTX1* mutations in HGBL we further identified recurrent mutations affecting *NCOR1* and others (**Supplementary Figure 5**) (14, 17). NOTCH signaling was thereby the predominant target of somatic mutation in HGBL-DH/TH, albeit with a quite heterogeneous mutational pattern affecting 35/47 patients with lesions in 28/71 genes **(Supplemental Figure 6A**). Most of these genomic aberrations had been previously reported to be gain-of-function mutations putatively resulting in constitutive NOTCH pathway activation in various types of predominantly germinal-center derived B-cell lymphoma. Several of these mutational hits including *NCOR1* and *DTX1* have been shown to herald adverse clinical outcome (34, 35). This remained the case when undertaking a differential downstream analysis within the cytogenetically defined subgroups (HGBL with *MYC* and *BCL2* aberrations, HGBL with *MYC* and *BCL6* aberrations and HGBL-TH), which was prompted by their significantly divergent distribution onto molecular clusters. Through this analysis a mutational signature became apparent that is additionally dominated by impairment of TP53 and MYC signaling in *BCL2* rearranged cases. *BCL6* rearranged cases lacked this enrichment, while HGBL-TH cases revealed intermediate mutational features. As another remarkable feature throughout all subgroups we observed alterations, putatively resulting in gain-of-function in IL6/JAK/STAT signaling in 74% of patients (**Supplementary Figure 5**). Activating mutations affecting *PIM1* and *SOCS1* were most frequently encountered in our case series and have been previously implicated in HGBL-DH/TH pathogenesis (17). These candidate driver genes are supplemented by mutations in *LTB* and *STAT3* (both previously identified in HIV-associated plasmablastic lymphoma) among others (36). Beyond this, we observed a relatively dispersed mutational pattern with putative driver events affecting 28 genes within the NOTCH-pathway **(Supplemental Figure 6A**).

In accordance with previous studies, we found mutations directly impacting NF-kB signaling in 62% of cases (**Supplementary Figure 5**) (37). While this was among the predominant pathways identified through our network propagation approach, mutations affecting the pathway were narrowly detectable with only *BCL2* harboring mutations in more than three patients (34%) followed by recurrent SNVs and indels in *PARP1* (6%) and *BIRC3* (4%) and *CARD11* (4%).

Following SNV and InDel evaluation with MUTSIGCV, a network propagation approach (**Figures 3B, C**) was employed on significantly mutated genes to delineate the functional implications of significant genetic events on neighboring genes. This further underscored the mutational impairment of the aforementioned pathways alongside WNT and PI3K signaling. These observations are in accordance with preliminary impressions derived from targeted sequencing studies, employing panel-based approaches (14, 17). In addition to the divergent results from our mutational pathway analysis, we identified an enrichment of E2F targets impacted by significantly mutated genes in both double- and triple-hit cases affected by *BCL6* rearrangements (**Figures 3D, E, F**).

### Survival analysis

Upon integrated analysis of molecular and clinical data we investigated genomic alterations present in > 15% of patients for their impact on overall survival (OS) and progression-free survival (PFS). Hereby we identified *ARID1A* mutations to be predictive of worse clinical outcome in our cohort (OS: p = 0.0049; PFS: 0.025). Subsequent Bonferroni correction for multiple testing was performed. Thus, we identified a significant impact of mutations affecting *ARID1A* which was maintained regarding OS when correction for multiple testing was applied while its primarily significant effect on PFS was reduced to a trend of borderline statistical significance (**Figure 5**). A Cox proportional hazard model revealed this effect to be independent of the established clinical IPI prognosticators (age, LDH, extra nodal manifestations, stage, and performance status; OS: p < 0.001; HR: 13.989; 95%CI: 3.362 – 58.205; PFS: p = 0.001; HR: 6.648; 95%CI: 2.098 – 21.061). Within the cytogenetically defined subgroups, we identified no alterations with independent impact on clinical outcome. However, a trend of borderline statistical significance towards inferior outcome in *MYC*/*BCL2* rearranged cases harboring *FOXO1* mutations was observed (**Supplementary Figure 7**).

**Figure 5.**
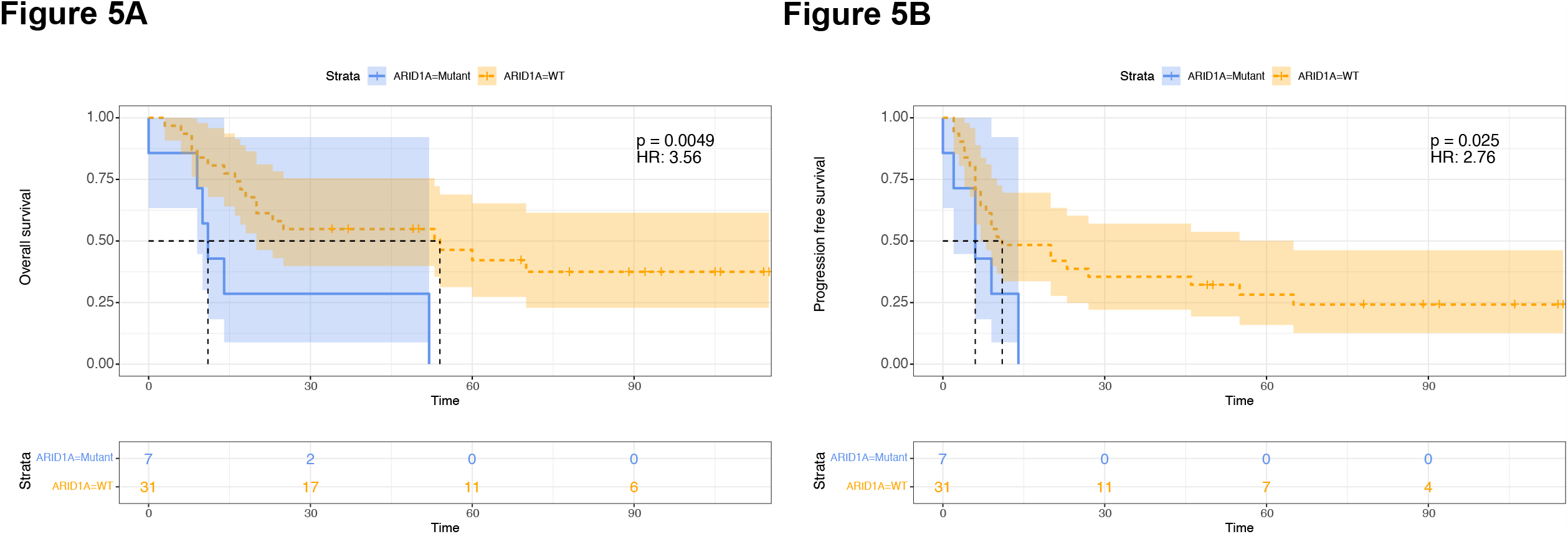
Overall (A) and progression-free survival (B) according to *ARID1A* mutational status. Numbers at risk alongside hazard ratios and p-values according to log-rank testing are provided. Subsequent Bonferroni correction for multiple testing (all genes with a mutational frequency > 15% were investigated) identified a significant impact of *ARID1A* mutations regarding OS while its primarily significant effect on PFS was reduced to a trend of borderline statistical significance.

## Discussion

Here we report on whole-exome sequencing data from an extensive cohort of HGBL-DH/TH tumors, which is to the best of our knowledge the hitherto largest cohort and most extensive molecular data set for this entity. Previous reports on HGBL-DH/TH were limited by low sample numbers and/or targeted sequencing approaches. Contrary to this, WES here allowed to systematically define recurrent mutations, predominant mutational signatures and SCNVs in their respective clinicopathological context from which we report three central observations.

Firstly, being the first exome-wide mutational investigation for this rare subtype of lymphoma, we identify a significant overlap of mutational drivers between HGBL-DH/TH and FL as well as GCB-type DLBCL (e.g., *TNFRSF14, EZH2* and *IRF4*) as its high-grade counterpart. Aggressive transformation was associated with the acquisition of mutations in *TP53*. Moreover, shared features, including *CCND3* and *CDKN2A* mutations underscore a close molecular relation between HGBL-DH/TH and BL (38, 39). This is additionally reflected in the enrichment of HGBL-DH/TH patients with Burkitt-like morphology for *CCND3* mutations. Further, we identify a number of significant mutational drivers not captured by previous, panel-based sequencing studies. Most frequently among these, we find *SI* mutations that have been previously implicated in CLL progression aa well as mutations in *POU2AF1*, which has been recently found to be an augmented target of mutations during aggressive transformation of FL to DLBCL (40, 41). Although *MYC* did not meet the predefined MUTSIGCV significance level in our study, we still observed mutations in 19% of cohort samples (**Supplementary Figure 8**), which is in agreement with previous panel-based studies (17).

Secondly, upon screening the mutational landscape in HGBL-DH/TH in comparison to the molecular clusters of DLBCL, proposed by Chapuy *et al*. and the LymphGen algorithm proposed by Wright *et al*., we unveil a striking overlap of *BCL2* rearranged cases with the C3/EZB cluster, which was previously shown to be enriched for *MYC* rearrangements and oncogenic drivers implicated in FL pathogenesis (22, 23). We argue that a predominant subset of HGBL-DH/TH most likely corresponds to these transformed FL. This offers a potential explanation for the inferior clinical outcome of C3 DLBCL patients, despite their GCB-phenotype, through an enrichment for *MYC* rearranged HGBL-DH/TH cases. Of note, we find the predominant impairment of TP53 and to a lesser extent MYC signaling in *BCL2* rearranged cases to be in keeping with a previous study on an independent set of HGBL-DH/TH, in which we found *TP53* mutations to be a recurrent feature of HGBL-DH with *BCL2*, but not *BCL6* rearrangements (12). Intriguingly, we further observed two molecular subtypes in *MYC/BCL6* only rearranged cases. While selected cases were categorized within the EZB cluster, several cases revealed an association with the BN2 cluster, potentially hinting at a *MYC*-driven high-grade transformation of a precursor lesion with an origin within the marginal zone, as previously described (23). In addition to these observations, we found triple-hit cases to reflect EZB lymphomas in the vast majority of cases, potentially hinting at *BCL6* rearrangements as late and non-defining events in HGBL-TH lymphomagenesis. Supporting this assumption, Pedrosa *et al*. have shown DLBCL with *BCL2* and *BCL6*, but without *MYC* rearrangements to be exclusively associated with the EZB cluster (42). Considering the significantly mutated genes, our observations underscore previous assumptions regarding a molecular divergence between *BCL2* and *BCL6* rearranged HGBL-DH (14, 17, 43). The predominant mutational distinction between these groups was the presumably FL-derived enrichment for *CREBBP* mutations in the *BCL2* rearranged subgroup. On an exome-wide level we observed an enrichment of the SBS6 signature (implicated in defective DNA mismatch repair) and a significantly diminished congruency with the C3/EZB DLBCL cluster in the *BCL6* rearranged subgroup. Of note, these findings fundamentally dispute the combined characterization in the current WHO classification, despite several shared clinical aspects common to all subtypes of HGBL-DH/TH (1, 2). Beyond *de novo* DLBCL with *BCL6* rearrangement, potential alternative explanations for this phenomenon include both clonal evolution and subsequent aggressive transformation from rare cases of *BCL6* rearranged marginal zone lymphomas alongside *BCL2* non-rearranged/*BCL6* rearranged FL, which were previously shown to be characterized by a heterogenous mutational landscape (44, 45). From our data, we further deduce an intermediate role for HGBL-TH, which may indicate two divergent paths of clonal evolution originating from a *BCL2* or a *BCL6* driven disease with subsequent acquisition of the alternative rearrangement. Lastly, we describe a pronounced mutational impairment of NOTCH, IL6/JAK/STAT and NFκB signaling pathways and recurrent oncogenetically relevant genes affected by SCNVs (including *MEF2B*, which was previously shown to be enriched in mutations/aberrations within the C3 DLBCL cluster) thereby systematically characterize the oncogenetic footprint of this subgroup of lymphoma. This is further combined with the identification of novel putative mutational drivers (e.g., *NCOR1, DTX1, LTB* and *STAT3*) alongside several previously established mutational hotspots in HGBL-DH/TH. Moreover, among these significantly mutated genes we describe *ARID1A* which emerges as a potential prognosticator of treatment response and outcome from our correlative assessment of clinical and molecular features of our present cohort, which was found to be independent from previously established clinical prognostic factors.

We acknowledge the shortcomings inherent to the retrospective design of the study alongside the limited availability of germline DNA for matched pair analysis. The latter aspect is reflected in a significantly elevated number of mutations in non-matched samples. This prompted us to limit our subsequent analysis to significantly mutated genes (except for *MYC* and *BCL2* mutations, which were additionally included based on their proven relevance in prior studies (14, 17, 22, 46) and thereby equalizing the abovementioned effect. Pairing of our WES-results with RNA-seq data, preferably in an extended, clinically annotated cohort, which was beyond the scope of the present study, would further deepen our molecular understanding of HGBL-DH/TH, especially regarding cases with prominent Burkitt or Burkitt-like morphology.

In summary, our identification of distinct mutational landscapes among HGBL-DH/TH, derived from an exome-wide sequencing approach shows both overlapping and distinctive features compared with germinal center derived lymphomas such as GCB-type DLBCL and low-grade FL as well as BL. Our work further underscores the developing notion of a recurrent pattern of mutational events driving a potentially unidentified preexisting FL into *MYC*-driven HGBL-DH/TH, offering insight into the molecular pathogenesis of this provisional entity. By refining the molecular taxonomy for aggressive, germinal-center derived B-cell lymphomas, these results call into question the current WHO classification system, especially regarding the status of *MYC*/*BCL6* rearranged HGBL.

## Data Availability

https://ega-archive.org/

## Acknowledgments

A.K. and H.B. acknowledge computational support from the OMICS compute cluster at the University of Lübeck. The research was supported by a grant to N.G. by the Stefan-Morsch-Foundation alongside infrastructural support. H.B. acknowledges funding by the Deutsche Forschungsgemeinschaft (DFG, German Research Foundation) under Germany’s Excellence Strategy— EXC 22167-390884018).

