## Supplementary Material for "Mutational landscape of high-grade B-cell lymphoma with *MYC-, BCL2* and/or *BCL6* rearrangements characterized by whole-exome sequencing"

**Supplement**

**Supplementary Materials and Methods**

**Clinicopathological characteristics of the study group**

Both conventional and immunohistochemical slides were reviewed by a panel of experienced hematopathologists in a multi-step process, where diagnosis was confirmed by two experienced hematopathologists (ACF, HM) in accordance with the current edition of the WHO classification of tumors of the hematopoietic and lymphoid tissues (1). From 80 cases meeting diagnostic and clinical criteria for HGBL-DH/TH with available biopsy specimen, 47 were selected for subsequent genomic analysis, based on tumor DNA quality and library preparation success.

Antibodies and positivity cutoffs employed in the current study are summarized in **Supplementary Table 1**. Fluorescence *in situ* hybridization (FisH) for *MYC* (including *MYC*-Ig fusion), *BCL2*, *BCL6* and chromogenic *in situ* hybridization for EBER were performed, as described (2, 3).

Clinical information was collected from the original files, and patients’ performance status (Eastern Cooperative Oncology Group [ECOG]), stage, treatment modalities, therapeutic response, pattern of relapse, baseline serum levels of lactate dehydrogenase (LDH), revised international prognostic index (R-IPI)(4) and information on survival were anonymously coded alongside hematopathological assessments. Extent of disease was routinely evaluated according to the Cotswold modifications of the Ann Arbor classification (5).

**Extraction of nucleic acids**

Tumor as well as germline DNA (when available from biopsies taken from non-involved sites; n = 7) was extracted from three FFPE tissue sections of 5µm thickness using Maxwell^®^ RSC DNA FFPE kit (Promega), according to the manufacturers’ instructions. Quality assessment and quantification was performed on an Agilent 2100 Bioanalyzer system (Agilent Technologies).

**Targeted next generation sequencing**

Library preparation was carried out according to manufacturers’ instructions and sequencing was performed on the Illumina MiSeq platform (Illumina, San Diego, California, USA) to a median depth of 3569x (s.d. ± 1635).

**Panel resequencing data analysis**

Resequencing data was processed as described above for whole-exome data, but the remove duplicates step was omitted. Variant calling was done using freebayes (v1.3.2-46-g2c1e395), variants were annotated using ANNOVAR and coverage for each variant was extracted using vcf-query (6). Afterwards, variants were filtered and only variants with a minimum coverage of 100, minimum variant allele frequency of 10% and population allele frequency < 0.001 in gnomAD or PopFreqMax database were kept for further analysis.

**Variant Calling in exome sequencing data**

Raw paired-end data (*fastq* format) was trimmed and quality filtered using fastp(7) (v0.20.0; minimum length 50bp, max. unqualified bases 30%, trim tail set to 1) and trimmed reads were mapped to GRCh37/hg19 using bwa mem (v0.7.15)(8). Resulting alignment files in *SAM* format were cleaned and sorted and converted into *BAM* format using Picard Tools (v2.18.4). Next, mate-pair information was fixed, duplicates were removed and base quality recalibration was performed using Picard Tools(9) and dbSNP v138. Single nucleotide variants (SNVs) and short insertions and deletions (indels) were identified following the best practices for somatic mutations calling provided by GATK(10) for version 4.1.7 or higher for matched normal and unmatched tumor samples. Briefly, GATKs Mutect2(11) (v4.1.7.0) algorithm was applied to all *BAM* files with gnomAD variants as germline resource and the b37 exome panel data as panel of normal. In cases, where matched normal tissue was available (Cases 1-7), Mutect2 was run in matched tumor-normal mode. Afterwards, FFPE read orientation artefacts were identified and removed according to GATKs guidelines and left-aligned filtered variants were annotated using ANNOVAR(12) (v2019Oct24). Coverage for reference and alternative alleles for each variant were extracted using vcf-query (VCFtools v0.1.13(13)). The top 20 frequently mutated genes (FLAGS(14)) were removed from further analysis. Somatic variants were filtered as follows: Minimum coverage of 40, minimum variant allele frequency of 10%, variant allele frequency < 0.001 in gnomAD or PopFreqMax database. To identify genes that are more often mutated than expected, MutSigCV (v1.41)(15) was applied and potential driver genes were identified using p < 0.001 and q < 0.1.

**Copy number aberrations**

The genomic landscape of HGBL-DH/TH was assessed for somatic copy number aberrations (SCNAs) by CONTROL-freec (v11.4)(16) using the tumor-only mode for samples without matched and normal tissue and in matched normal-tumor mode for samples with normal tissue available (cases 1-7). The output (ratio and reads per called window) was converted to run GISTIC2.0 (v2.0.23)(17) excluding CNA calls from chromosomes X and Y and excluding known common CNAs using Broad Institute’s panel of normal (ftp://ftp.broadinstitute.org/pub/GISTIC2.0/hg19_support). GISTIC2.0 analysis was performed using default parameters.

**Mutational significance and deleteriousness, network propagation, gene set enrichment and mutational cluster analysis**

The effect of strong deleterious effects (CADD1.3 phred score > 20) was assessed per sample using a network propagation approach(18) applying a regularized Laplacian kernel based on STRINGdb v11(19) protein-protein interaction network (diffuStats v1.8.0(20)). Genes affected by strongly deleterious mutations were set to 1, whereas non-mutated genes were set to 0 to model the behavior of the mutation and network diffusion was performed using a parametric method with statistical normalization (*z*-scores).

The effect of potential driver genes identified by MutSigCV on neighboring genes was assessed using a network propagation approach (as described above). Resulting *z*-scores were used as pre-ranked input for a rank-MANOVA based statistical approach to detect enriched gene sets (mitch R packages)(24, 25). Gene set enrichment was performed against HALLMARK gene sets and the NF-kB signaling pathway (genes were retrieved from KEGG; entry ID hsa04064). In addition, the acquired genomic data were processed through the LymphGen algorithm and the mutational patterns sequentially underwent cluster analysis and were subsequently screened manually for an enrichment in overlapping aberrations with the molecular clusters proposed by Chapuy *et al*. (26, 27). Further, a logistic regression framework was employed in order to test for significantly different numbers of mutated genes in a given cluster between HGBL harboring different cytogenetic constellations.

**Statistical Analyses**

If not stated differently, all statistical analyses were performed using R (v4.1.0) and tidyverse (v1.3.1)(28) for data handling. Filtering of genomic regions was performed using GenomicRanges (v1.44.0)(29) and maftools (v2.8.0)(30) were used to visualize the data. The number of somatic signatures and the contribution of each signature to each case was estimated based on the YAPSA (v1.18.0) package with calculation of correction factors for WES to avoid biases in the k-mer distribution introduced with the exome capture kit. Progression-free survival and overall survival (PFS, OS) were calculated from the date of diagnosis and censored at last clinical contact. Survival (PFS and OS) according to potential prognostic factors was estimated by means of the Kaplan–Meier method and univariate log-rank test. Survival analysis was carried out employing the R packages survival (v3.2-11) and survminer (v0.4.9).

**Supplementary Table 1.** Antibodies and positivity cutoffs employed in the current study

| **Antibody** | **Supplier** | **Clone** | **Positivity cutoff** |
| --- | --- | --- | --- |
| Bcl2  | Lab Vision | 100/D5 | 30% |
| Bcl6 | Dako | BG-B6p | 30% |
| CD10 | Menarini | 56C6 | 30% |
| CD20 | Dako | L26 | - |
| CD30 | Dako | BerH2 | 10% |
| CD38 | Leica Biosystems | SPC32 | - |
| CD138 | Leica Biosystems | MI15 | - |
| CD56 | Leica Biosystems | CD564 | 10% |
| Kappa | Leica Biosystems | CH15 | - |
| Lambda | Leica Biosystems | SHL53 | - |
| MUM-1 (Irf4) | Dako | Mum 1P | 30% |
| Ki-67 | Dako | Mib-1 | - |

**Supplementary Table 2.** Gene list for the custom AmpliSeq panel (Thermo Fisher Scientific, Waltham, Massachusetts, USA) for targeted amplicon sequencing – see separate .xlsx file

**Supplementary Table 3.** Genes found to be significantly mutated in HGBL-DH/TH according to the MutSig2CV algorithm – see separate .xlsx file.

**Supplementary Table 4.** All variants described by WES – see separate .xlsx file**.**

**Supplementary Table 5.** All variants described by panel based NGS – see separate .xlsx file.

**Supplementary Table 6.** Association between HGBL-DH/TH cases and mutational signatures derived from WES data – see separate .xlsx file.

**Supplementary Table 7.** LymphGen predictions for each case – see separate .xlsx file.

**Supplementary Table 8.** A logistic regression framework assessing the number of C3 subgroup mutations present in *MYC* rearranged with additional rearrangements of *BCL2*, *BCL6* or both.

|  | **Estimate** | **Std.Error** | **t-value** | **p-value** | **CI_Lower** | **CI_Upper** | **DF** |
| --- | --- | --- | --- | --- | --- | --- | --- |
| **(Intercept)** | 2.5714 | 0.2893 | 8.89 | 2.21*10^-11^ | 1.988 | 3.1544 | 44 |
| **BCL2/6** | -0.6714 | 0.4004 | -1.677 | 1.01*10^-01^ | -1.478 | 0.1356 | 44 |
| **BCL6** | -1.7589 | 0.3681 | -4.778 | 2.00*10^-05^ | -2.501 | -1.0171 |  |
| Multiple R-squared: 0.3407, Adjusted R-squared: 0.3108  F-statistic: 12.21 on 2 and 44 DF, p-value: 6.059e-05   CI = 95% confidence interval, Std. Error = standard error. DF = degrees of freedom. | | | | | | | |

**Supplementary Figure legends**

**Supplementary Figure 1.** Distribution of mutations within selected, significantly mutated genes in the format of lollipop plots

**Supplementary Figure 2.** Panel (A) depicts the profiling of mutational signatures driving HGBL-DH/TH revealed a predominance of the SBS5 signature alongside the emphasized occurrence of the SBS6 signature (implicated in defective DNA mismatch repair) in patients with *BCL6* rearrangements. Additionally, covariates are shown above the plot for each sample. Panel (B) displays the proportion of the SBS6 signature according to *BCL6* FISH status, showing an elevated frequency in *BCL6* rearranged cases.

**Supplementary Figure 3.** Allocation of HGBL-DH/TH samples unto the molecular subgroups/clusters of DLBCL, according to Chapuy *et al.* based on their mutational signature(22). Additionally, covariates are shown above the plot for each sample.

**Supplementary Figure 4.** Concordance of *BCL2* and *BCL6* rearranged cases with the C3 DLBCL mutational Cluster according to Chapuy *et al.* by means of a logistic regression model; ^***^ denotes p < 0.001 (see **Supplementary Table 8** for details).

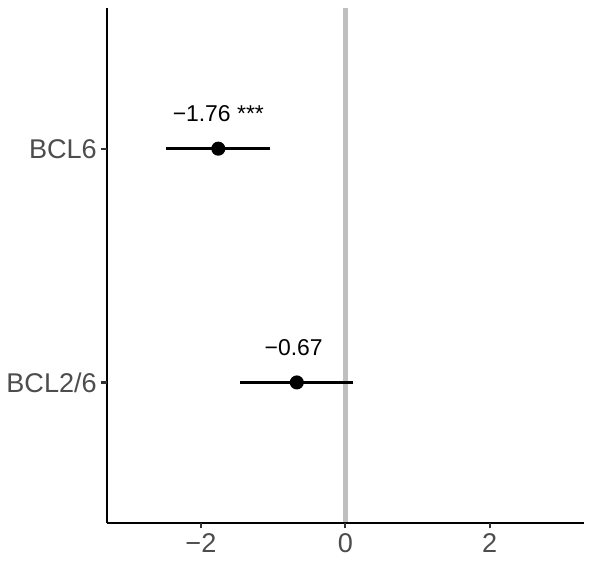

**Supplementary Figure 5.** Oncoplot depicting distribution of mutations unto the genes of NOTCH, IL6/JAK/STAT and NF-kB signaling pathways. Median of variant allele frequency per gene across mutated samples is shown in the left bar plot; the bar plot on the right-hand side shows the number of mutated samples per gene and pathway. Subtypes were inferred applying the LymphGen algorithm.

**Supplementary Figure 6.** Distribution of mutations unto the different signaling pathways for the HGBL-DH/TH data set (A), the *BCL2* subgroup (B), *BCL6* subgroup (C), and the triple hit subgroup (D).

**Supplementary Figure 7.** Overall (A) and progression-free survival (B) according to *FOXO1* mutational status.

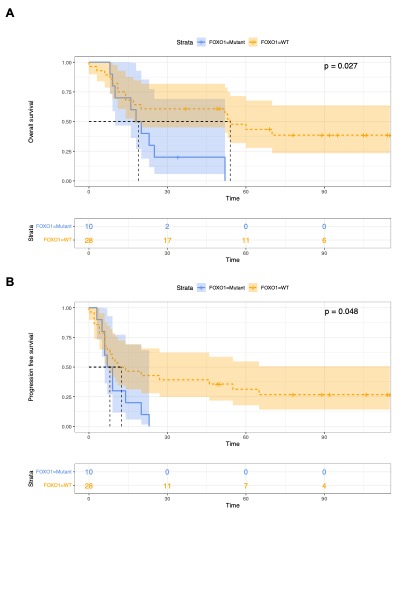

**Supplementary Figure 8.** Oncoplot depicting the mutational spectrum encountered in HGBL-DH/TH prior to filtering through the MutSigCV algorithm.
