## Supplementary Figure 1 for "Mutational landscape of high-grade B-cell lymphoma with *MYC-, BCL2* and/or *BCL6* rearrangements characterized by whole-exome sequencing"

*CCND3* : [Somatic Mutation Rate: 25.53%]
NM_001760

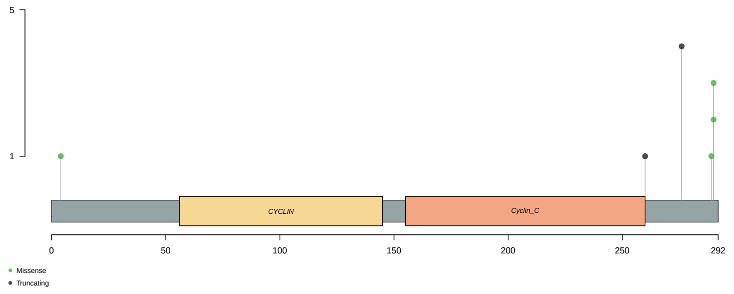


*CD79B* : [Somatic Mutation Rate: 8.51%]
NM_001039933

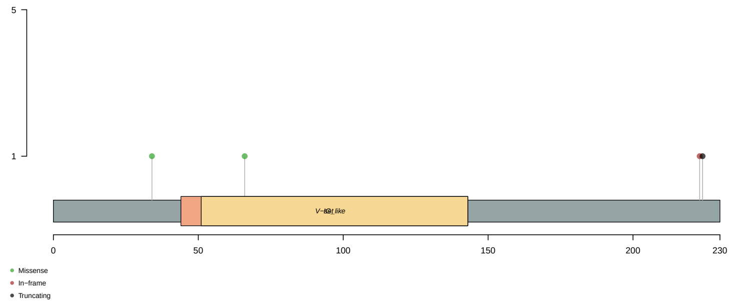


*CDKN2A* : [Somatic Mutation Rate: 10.64%]
NM_001195132

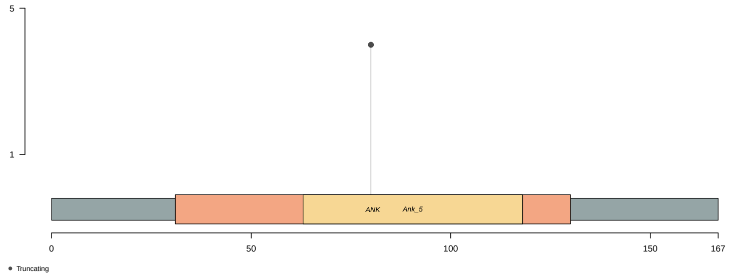


*GULP1* : [Somatic Mutation Rate: 8.51%]
NM_016315

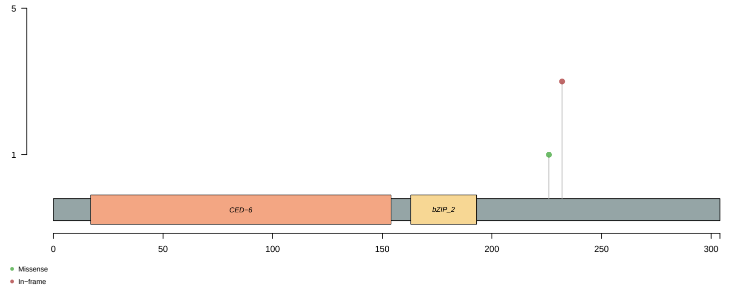


*IRF8* : [Somatic Mutation Rate: 12.77%]
NM_002163

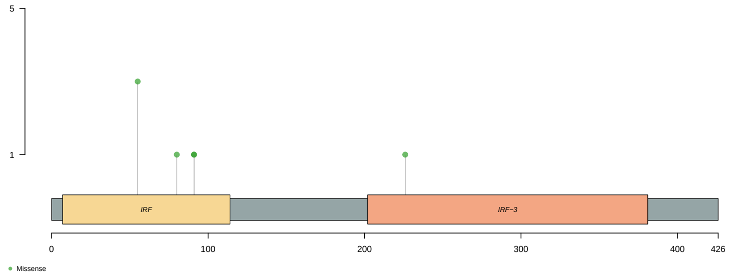


*KMT2D* : [Somatic Mutation Rate: 36.17%]
NM_003482

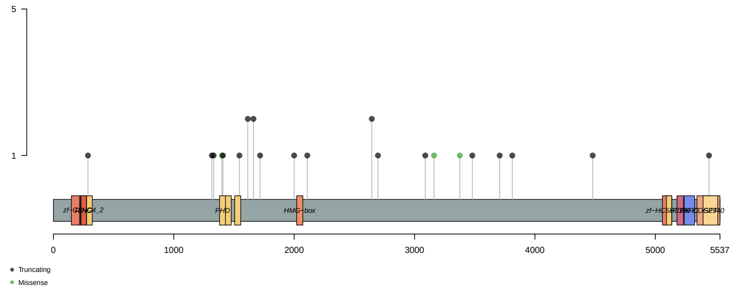


*POU2AF1* : [Somatic Mutation Rate: 8.51%]
NM_006235

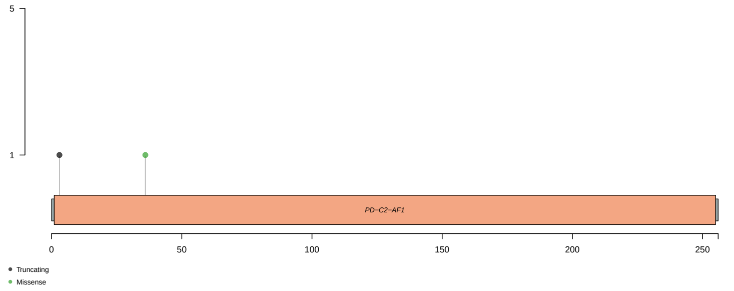


*RUFY2* : [Somatic Mutation Rate: 10.64%]
NM_017987

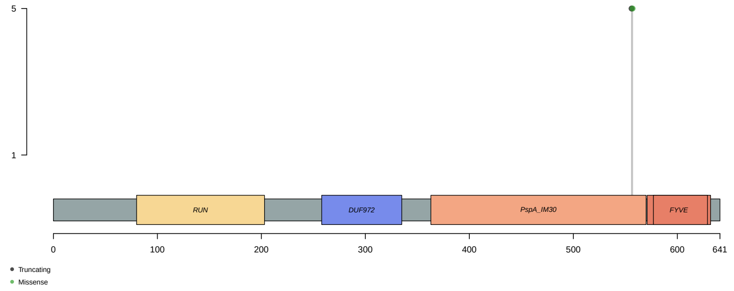


*SI* : [Somatic Mutation Rate: 21.28%]
NM_001041

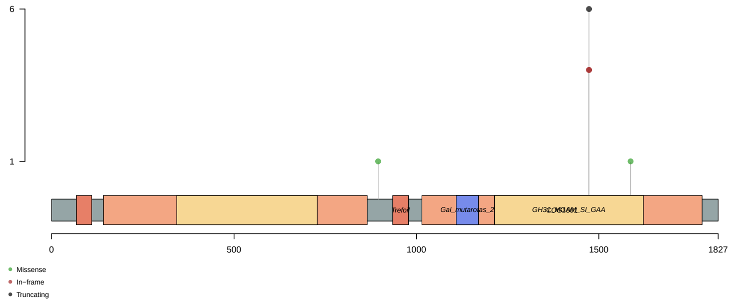


*TNFRSF14* : [Somatic Mutation Rate: 14.89%]
NM_003820

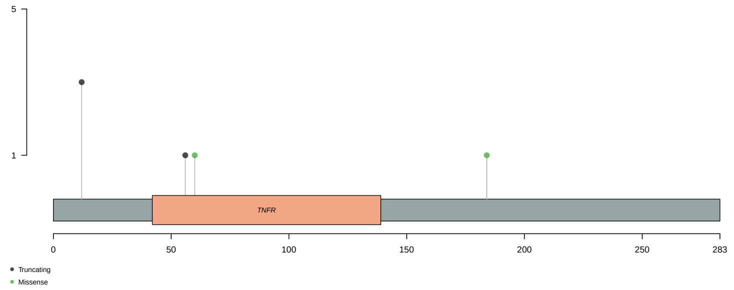


*TP53* : [Somatic Mutation Rate: 19.15%]
NM_000546

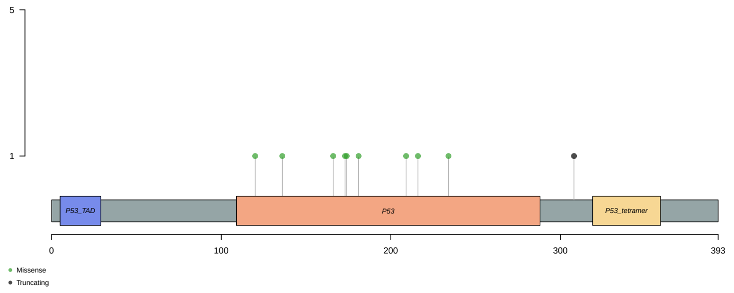
