## Supplementary figures and images for "Mutational landscape of high-grade B-cell lymphoma with *MYC-, BCL2* and/or *BCL6* rearrangements characterized by whole-exome sequencing"

### Supplementary Figure 2

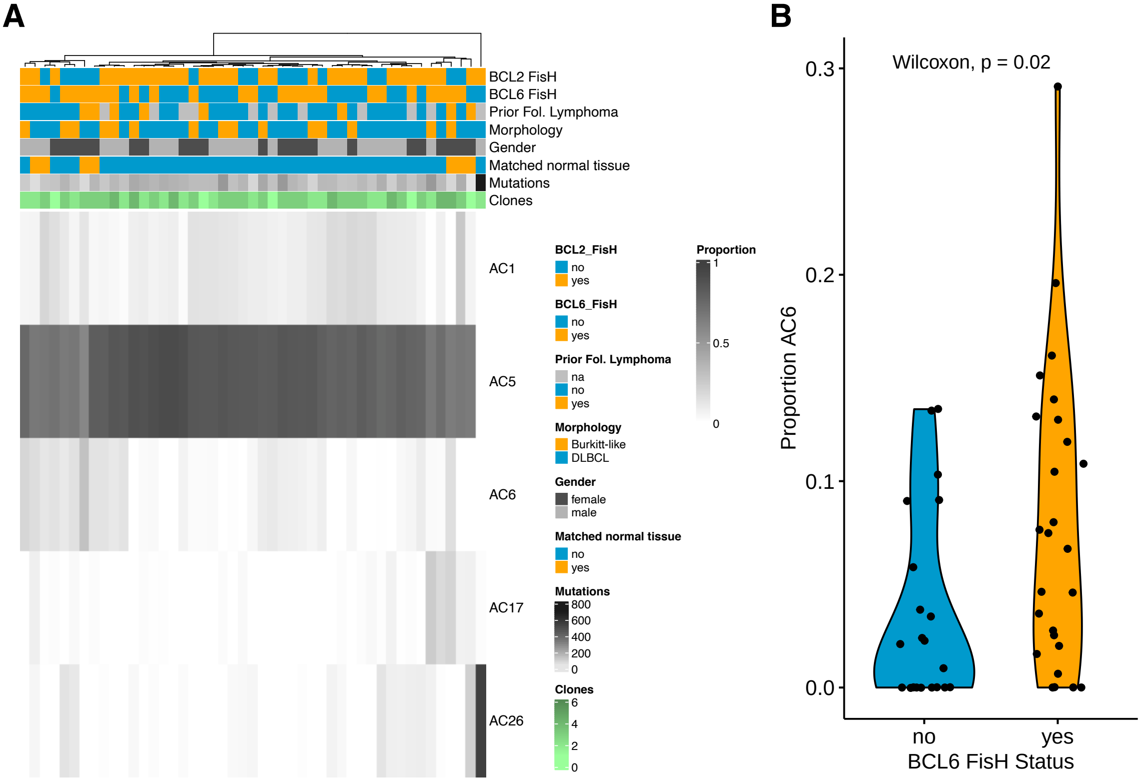

### Supplementary Figure 3

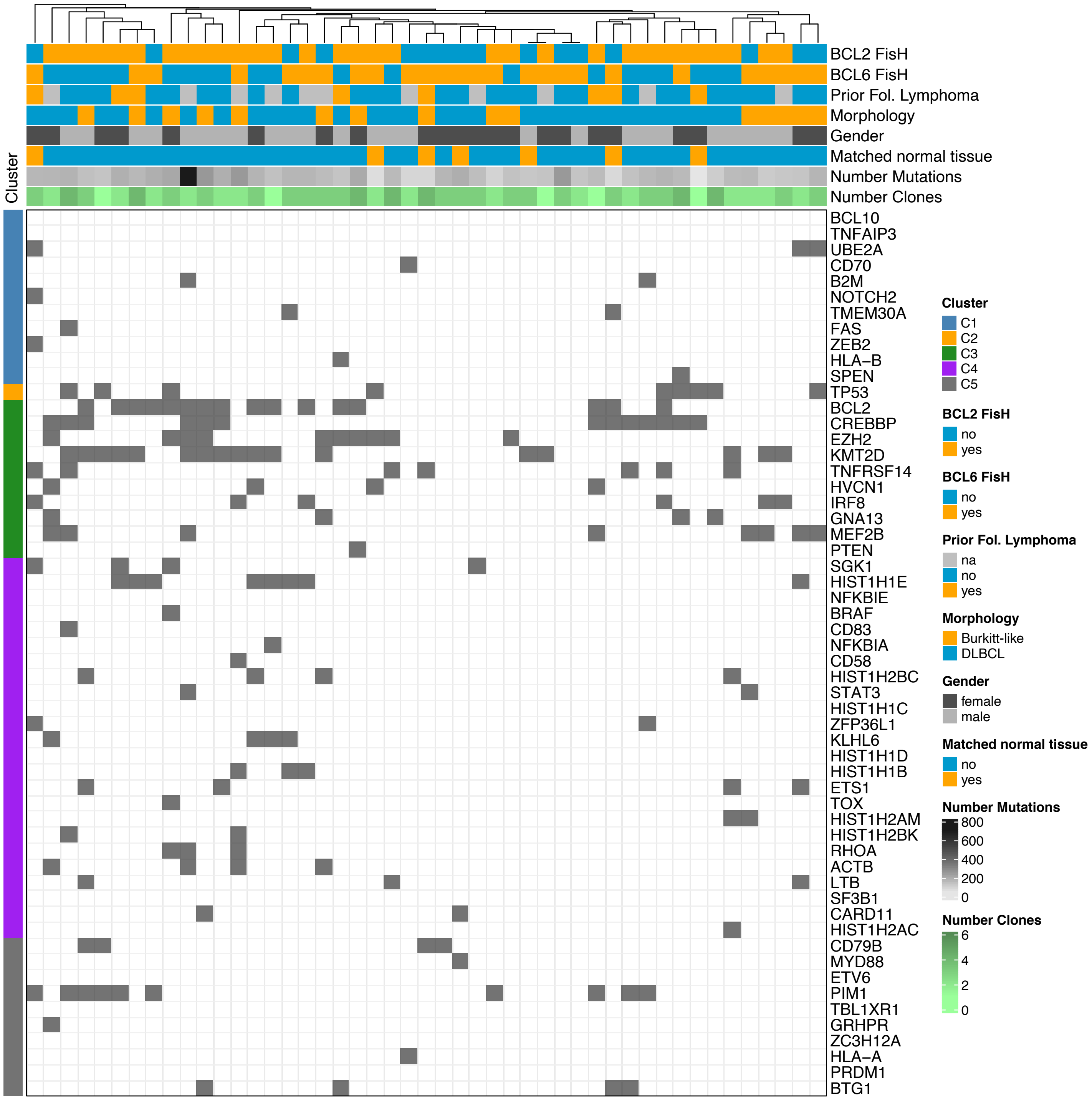

### Supplementary Figure 4

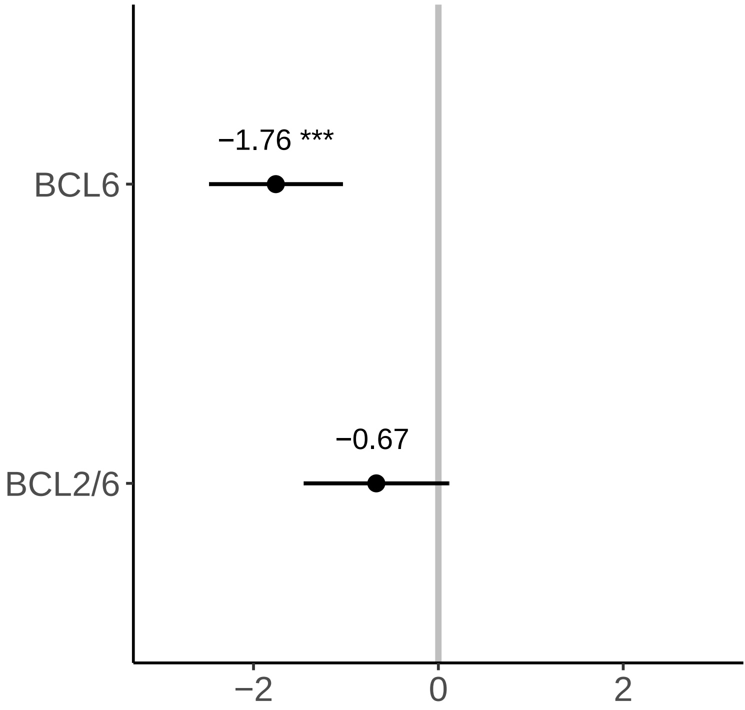

### Supplementary Figure 5

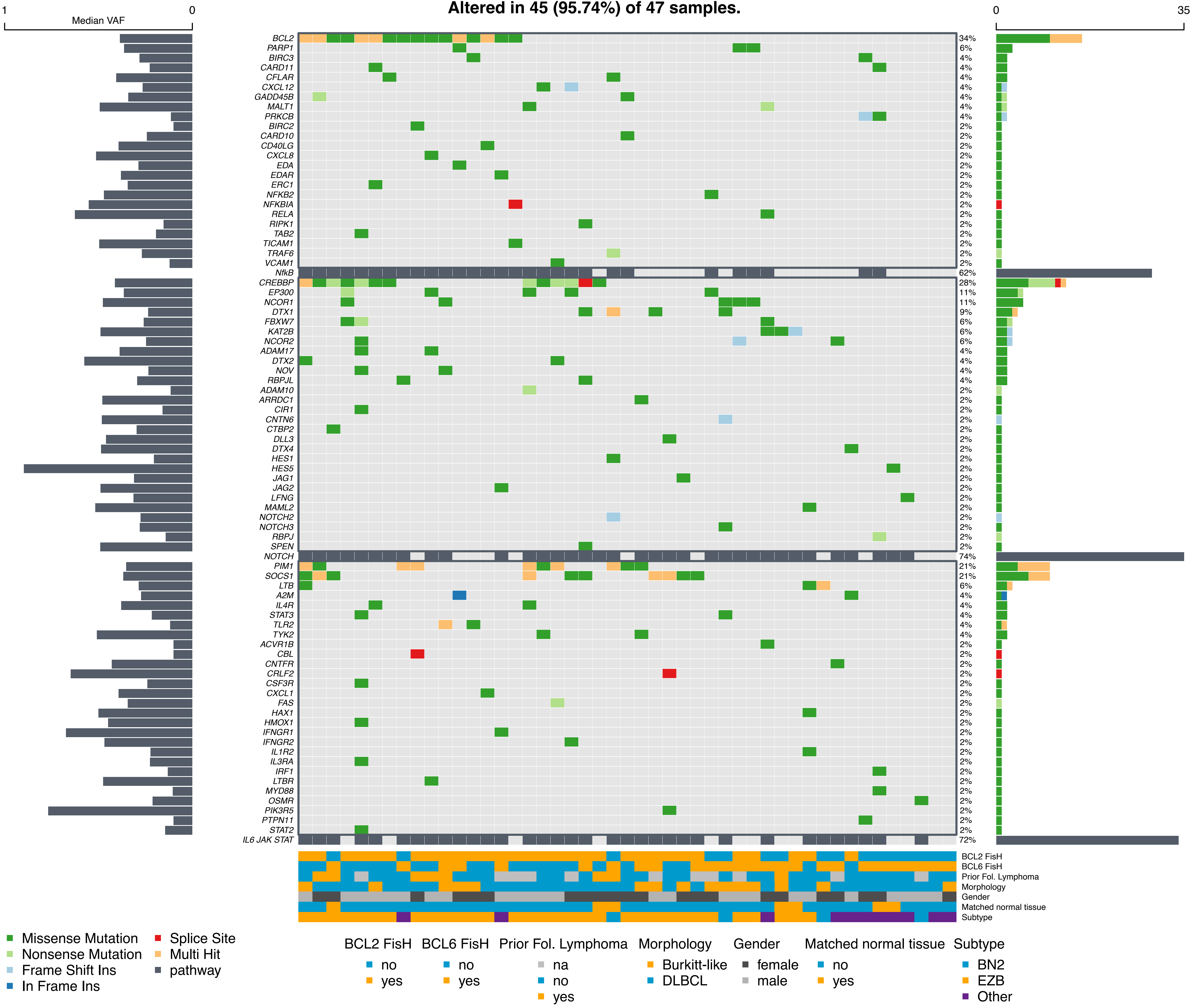

### Supplementary Figure 6

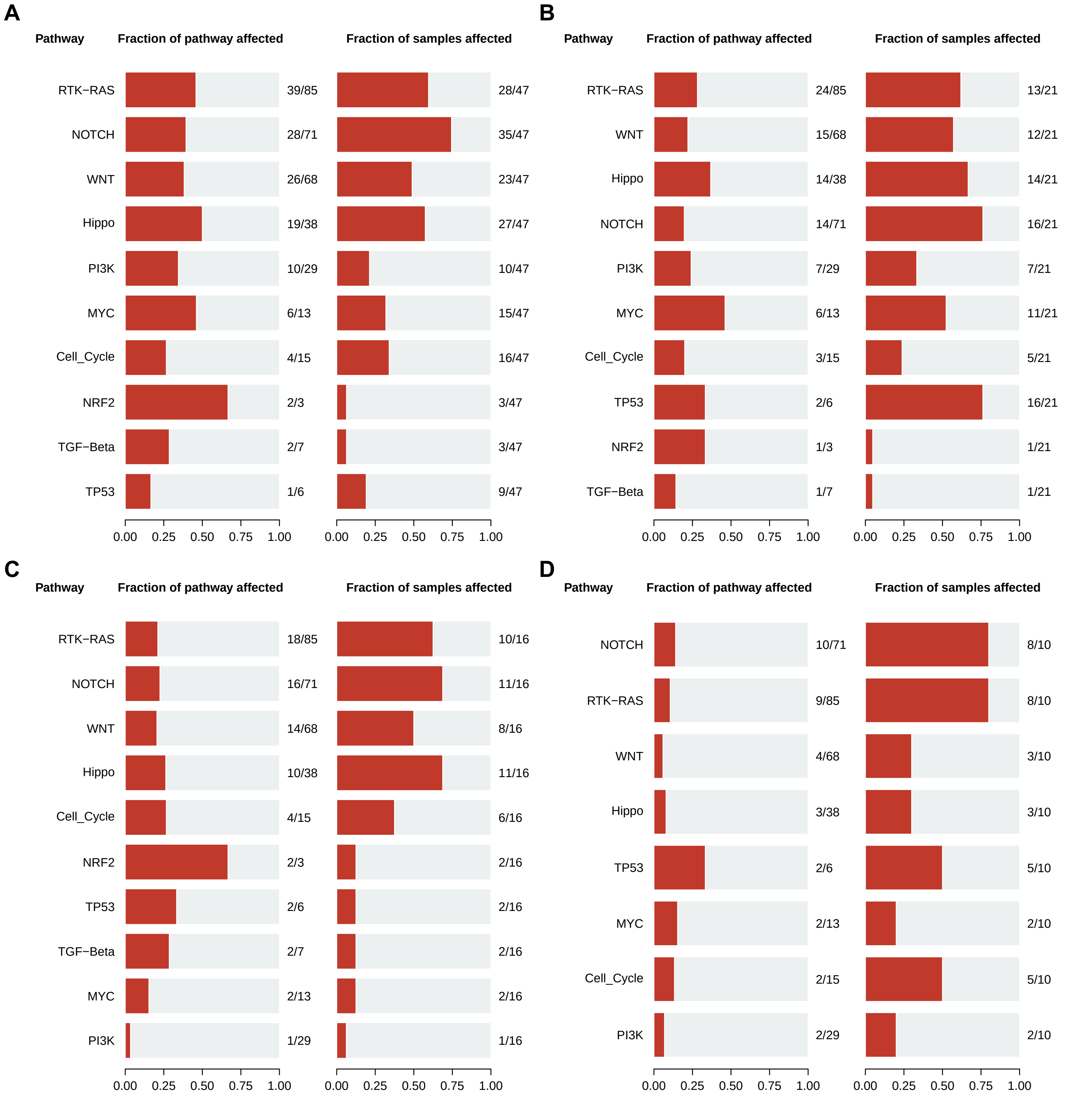

### Supplementary Figure 7

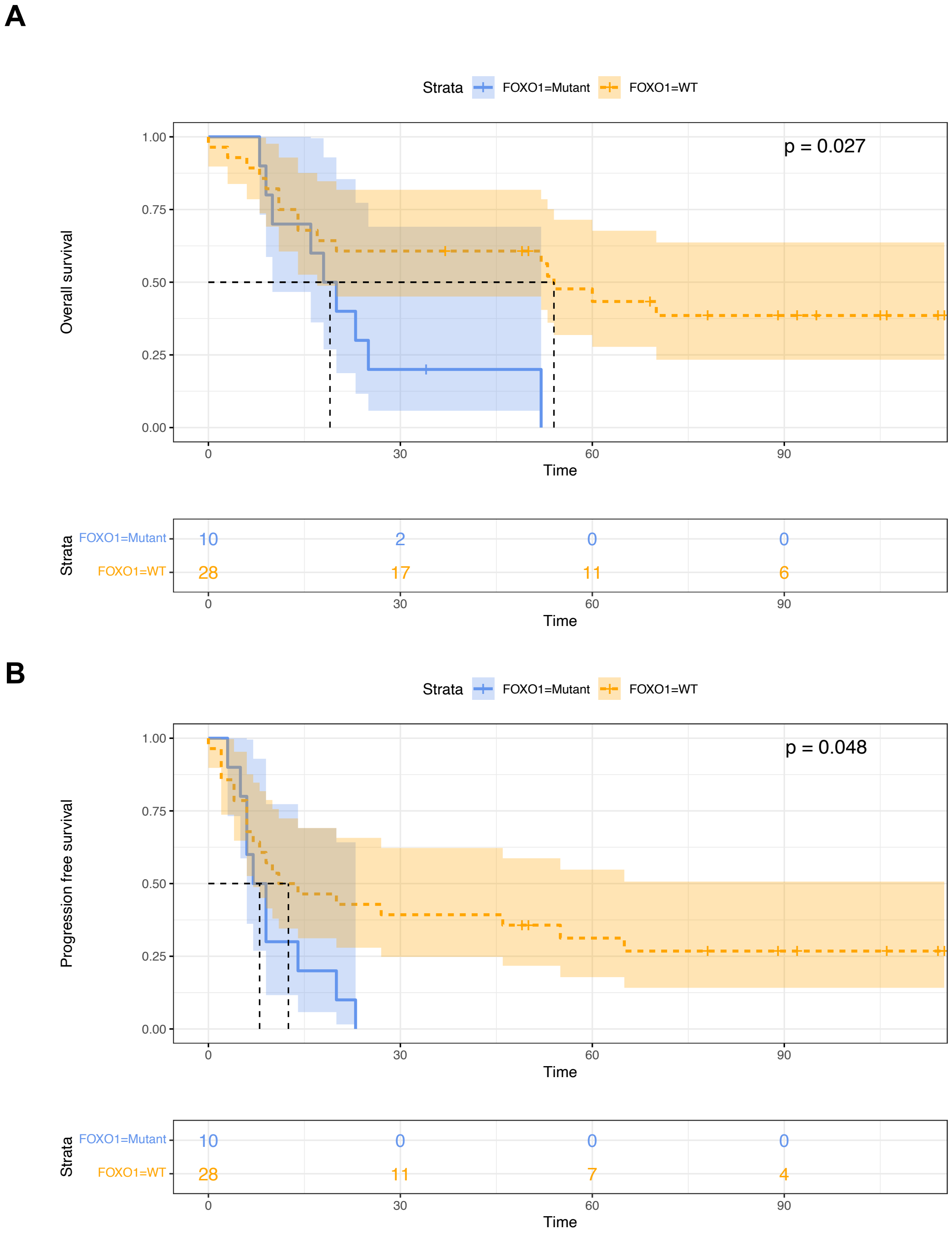

### Supplementary Figure 8

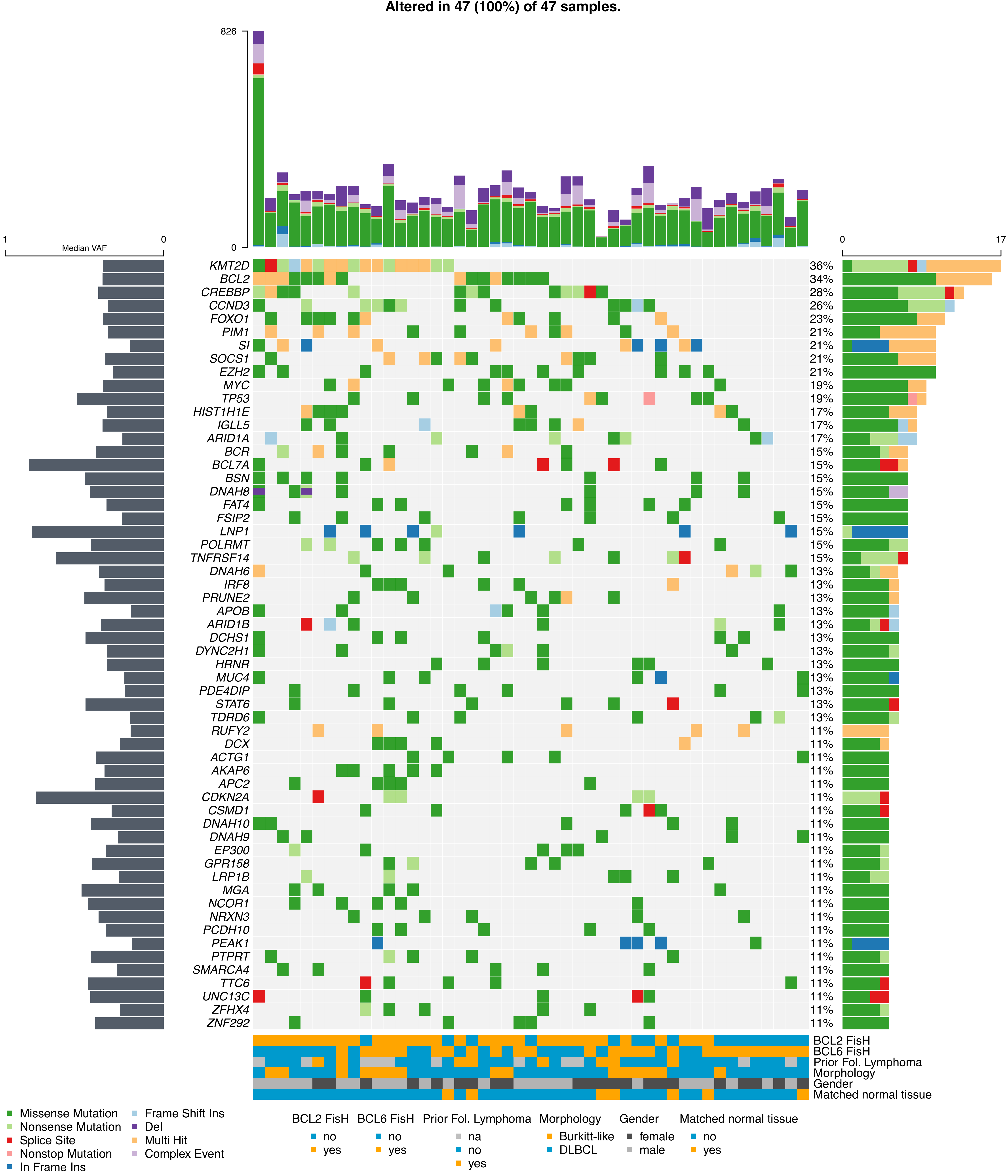
